# Pathways of health care for people living with multimorbidity in two southern African countries

**DOI:** 10.1101/2025.08.01.25332721

**Authors:** Gift Treighcy Banda-Mtaula, Mtisunge Joshua Gondwe, Nateiya M. Yongolo, Rashida A. Ferrand, Stephen Kasenda, Sara Lowe, Brown David Khongo, Naomi S. Levitt, Beatrice Matanje, Charlotte Taderera, Justin Dixon, Celia L. Gregson, Felix Limbani

## Abstract

Multimorbidity, the presence of multiple chronic conditions in one person, is a growing global health concern. Integration of chronic care services is urgently needed, especially in low-resource settings including in Southern Africa, where care has been fragmented by vertical and siloed disease approaches. Many countries share similar challenges to integration, presenting rich opportunities for shared learning. Yet, rarely are these opportunities capitalised upon, in part because of a lack of systematic knowledge about the similarities and differences in the health system contexts, challenges and current progress towards integration. As part of an inter-country collaboration, we sought to answer the questions: What are the common and distinct characteristics of the care pathways for people living with multimorbidity in Malawi and Zimbabwe, and the opportunities and challenges that emerge through such a country-level comparison? We used an iterative, qualitative research design that involved a desk review of relevant indicators, policies and strategies; key informant interviews, collaborative workshops, and the development of case studies of service integration in practice. Thematic analysis and comparison of challenges of integration across different levels of care revealed uneven funding for different diseases, a lack of both ‘vertical’ and ‘horizontal’ integration, frequent stockouts of drugs and diagnostic equipment, especially for noncommunicable diseases (NCDs), and inadequate training and support for clinicians. In both countries, progress towards decentralising and integrating chronic disease care at national level, has occurred through inclusion of specific NCDs into HIV programmes. This is prone to leave out comprehensive chronic care for people that are not living with HIV and reproduces verticalised programming. We suggest that a promising avenue for wider scale-up of decentralised, non-HIV-dependent integrated care lies in the expansion of an Integrated Chronic Care Clinic (IC3) model that provides comprehensive health system integration for all chronic diseases. Further cross-country learning and feasibility assessment is needed to advance this model.

## 1. Introduction

Multimorbidity, when two or more chronic conditions coexist in one person, is a global health concern.(1) Multimorbidity predisposes to loss of function, a lower health-related quality of life (HRQoL), polypharmacy (the use of many medications), and high socioeconomic costs.(4–6) People with multimorbidity are more likely to be hospitalized, remain longer in hospital, and die early than those with a single chronic disease.(7, 8) These associated factors raise healthcare utilisation costs, particularly in already financially constrained public health systems in Low and Middle Income Countries (LMICs).(9)

Patients with multimorbidity require integrated, person-centred approaches to optimise what is often complex care.(10) However, clinical guidelines, health management strategies, and wider health system architectures continue to prioritise the treatment of individuals with distinct diseases; i.e., the ‘disease silo’ model.(11, 12) Fragmentation of health systems is, while a challenge worldwide, especially stark in LMICs, including in sub-Saharan Africa, due to the dominance of ‘vertical’ programming (including funding) and historic biases towards acute, reactive care.

In response, several countries have taken steps to reduce disparities in care for NCDs and HIV by decentralising NCD care, as HIV care models have done previously, and by beginning to integrate chronic care services (e.g., hypertension and diabetes services). Initially, the focus was on adding NCD prevention and control into HIV care delivery.(13, 14) However, a study from Malawi, South Africa and Kenya demonstrated considerable variation in policy approaches to integrated HIV-NCD care across countries.(15) Increasingly, efforts have been made to expand integrated chronic care to all, irrespective of HIV status. While there are promising models that have been developed and implemented in Malawi, South Africa, Uganda, Tanzania, and Kenya, among others, this approach remains at an early stage and extremely challenged given entrenched legacies of vertical programming and enduring resource scarcity.(16)

Given the urgent need multimorbidity presents, and the similar challenges faced by many countries in the region, there is great potential for cross-country learning and research to advance integrated care approaches at national and regional levels. To date, however, these opportunities have not been optimally capitalised upon, in part because of a lack of systematic knowledge about the similarities and differences in the health system contexts, challenges and current progress towards integration. Through a comparative study of care pathways for people with multimorbidity in two countries in southern Africa, Malawi and Zimbabwe, we aimed to understand the similarities and differences in these pathways in the two settings, the facilitators and barriers to access to care (over and above those for single chronic conditions), and the extent to which the countries are moving towards integrated chronic care. Understanding is intended to reveal key opportunities for cross-country learning, research and interventions, and ultimately enhance a more integrated response to multimorbidity within and across these two settings.

## 2. Methods

### 2.1 Study design

This study was initially conceptualised during a regional workshop that drew together interdisciplinary expertise representing 10 African countries including Malawi and Zimbabwe, to devise shared concepts and priority areas for addressing multimorbidity in the region. Towards the end of the workshop, contributors self-organised into several working groups to consider opportunities for further investigation of various themes that emerged from proceedings. The broad area of focus for this working group was “health systems and care models”. The core themes raised for this workstream have been summarised in Table 2 of our previous publication.(17) The working group had a series of online meetings to reach consensus on the research question and aims, and the countries selected for inclusion.(18)

In carrying forward this theme, the working group devised a qualitative study design that leveraged existing research projects to facilitate cross-country research. The design consisted of a narrative desk review of policy documents and indicators related to the healthcare economies of Zimbabwe and Malawi, interviews with key stakeholders, collaborative workshops to interpret and refine findings, and the production of case studies exemplifying the strengths and weaknesses in multimorbidity care.

This research was embedded within larger studies namely:, Multilink in Malawi and KnowM in Zimbabwe. The Multilink Consortium is a multi-country study that aims to design and test a system which identifies patients suffering from multimorbidity (defined as at least two of hypertension, diabetes, HIV and chronic renal failure) during emergency assessments in sub-Saharan African hospitals, optimise emergency treatment, and ensure post-discharge linkage to appropriate care.(19) The KnowM study aimed to develop an interdisciplinary conceptual framework and research agenda to effectively respond to multimorbidity in sub-Saharan Africa.(20) While having distinct overall aims, both had embedded qualitative components with enough flexibility for a broadly similar approach across the two countries to answer our shared research question.

### 2.2 Desk review and data collection tools

A narrative desk review first aimed to provide a broad, formative understanding of care pathways in Zimbabwe and Malawi. A Google Search was conducted to retrieve policy documentation about the healthcare pathways for people living with multimorbidity to contextualize the interviewees’ information and better understand the main policy contents and pathways. Search terms included “chronic diseases” “non-communicable diseases,” “multimorbidity”, and the names of specific conditions (e.g. hypertension, diabetes, HIV, TB, etc.). In addition, the Malawi and Zimbabwe official Government websites, and those of relevant local and international technical partners were searched for key policy and strategy documents, and health economy information. The search period was from January 1st, 2023, to March 2025 when the manuscript draft was completed. S1 Table shows the reviewed documents from both countries.

Based on these reviews and the existing knowledge among the research team of the respective healthcare systems in Zimbabwe and Malawi, we developed a template core healthcare pathway for people with multimorbidity(S1 Figure) to guide stakeholder interviews and workshops (described below). These care pathways were iteratively developed and refined in each country, before being merged into one care pathway commensurate across both. In parallel, semi-structured interview guides were developed (in Malawi) or adapted from existing ones (in Zimbabwe) to ask about where people with multimorbidity (PwMM) source care, the accuracy of the drafted care pathway, and additional health providers, non-governmental organisations (NGOs), community support organisations (CSOs) and other partners who potentially contribute to the pathways. A table template was devised to describe service levels (S2 Table), with 5 sections including (i) service level/provider; (ii) service definition; (iii) service availability - nationally, regionally, and locally; (iv) challenges/barriers and (v) facilitators/solutions to service delivery.

### 2.3 Stakeholders

Using purposive and snowball sampling methods, we selected a sample of high-level stakeholders, including Ministry of Health representatives and technical partners actively involved in policy, planning, implementation, clinical care, and/or monitoring, with a focus on chronic disease and multimorbidity. These were identified based on reviewed policy documents (S1 Table), country-specific knowledge of the policy landscape, and from informal conversations with prospective interviewees. Engagement methods varied by country, given contextual differences and the studies within which each was embedded, but both sought to include the most relevant and diverse group of stakeholders possible, who had the greatest role in policy design and/or implementation. As we proceeded, new names were elicited and were included in the list of potential stakeholders. We finally interviewed ten stakeholders in Malawi and nine in Zimbabwe.

### 2.4 Procedures

Data were collected through in-depth interviews, followed by stakeholder engagement workshops. Interviews were facilitated by two authors (conducted in English either face-to-face or by video call). In both countries, interviews were audio recorded after obtaining stakeholders’ consent, or if stakeholders preferred, detailed interview notes were taken instead. The average interview duration was 40 minutes (range: 20 mins - 1 hour). Half-day workshops were conducted face-to-face in both Malawi and Zimbabwe, with detailed meeting notes taken of proceedings. Within these broad parameters, data collection was tailored to the country and research context.

In Malawi, 10 interviews with policymakers were conducted between 30-08-2023 and 11-09-2023. Interviews were guided by questions to explore policy makers’ perceptions of patient care pathways and known barriers and facilitators. Policymakers were from local government, Ministry, and funding bodies. The interviewees’ responses were then used to provisionally define the patient care pathway. The pathway was then refined through discussions with the NCD technical working group members at a workshop in September 2023 (attendees n=23). The workshop conducted with the technical working group had representatives from CSOs, Ministry directorates, research institutions, academicians, clinicians and funders. Stakeholders were asked to comment on the drafted pathway based on the interview policymakers’ opinion. Stakeholders refined various aspects of the care pathway based on their technical expertise and experiences and discussed barriers and facilitators to accessing care that emerged from the interviews.

In Zimbabwe, high-level findings from the broader KnowM study (described in detail elsewhere) (21) were used to provisionally define the care pathway and care provider table. As in Malawi, these were then refined through the in-depth interviews conducted with Ministry stakeholders and technical partners at national and sub-national level between 01-09-2022 and 31-12-2023.

Interviewees included national-level stakeholders representing departments for the administration of NCDs, HIV and TB (n=4) as well as provincial and district level administration (n=3), and international health organisations (n=2). As part of these interviews, stakeholders were asked about the care delivery pathway and challenges and facilitators for multimorbidity care, tailored to their specific areas of knowledge and expertise. For some respondents, this meant helping to refine and discuss particular aspects of the pathway, whereas others were in a better position to comment on the overall structure of the health service. The care pathway was finally presented as part of a collaborative workshop in Harare (attendees n=37). Stakeholders at the workshop were asked to comment on the pathway as well as challenges and limitations in the provision of services at different levels of care.

#### 2.4.1 Case study design

Complementing the findings from interviews and workshops, a case study was drafted from each country to demonstrate current developments in integrated chronic care and to share best practices for potential replication. The case studies reflected a facility or programme that was identified through conversations with informants during interviews and consultation workshops. Case studies were produced in a collaborative manner with those involved in the facilities/programmes, seeking to describe the current scenario with their successes and any experienced or potential challenges, as well as potential for scale-up

### 2.5 Data management and analysis

Data analysis was an ongoing, iterative process that folded new sources of information into ongoing research activities. Findings from the review of policy and health economy were summarised and cross-tabulated for analysis meetings among the research team, which fed into the development of the care pathway and interview guides. In-depth interviews were transcribed and entered into NVIVO 12. GTB, MG and JD coded the transcripts individually, and thematic analysis using grounded theory, with preliminary results used to refine the care pathway in each country, populate the comparison table, and feed into the discussions during collaborative workshops. The cross-country analysis was conducted through monthly meetings among the writing group which involved comparing and contrasting our model/tables as they evolved, which were used to devise core cross-cutting themes and finally to develop our main arguments and recommendations.

### 2.6 Ethical considerations

In Malawi, the College of Medicine Research & Ethics committee (COMREC) REF P.11/21/3462 approved the study (under Multilink). In Zimbabwe, the Medical Research Council of Zimbabwe (MRCZ/A/2842), Joint Research Ethics Committee of the Parirenyatwa Group of Hospitals and University of Zimbabwe Faculty of Medicine and Health Sciences (386/2021), the City of Harare, and the London School of Hygiene and Tropical Medicine (26469) approved the study (under KnowM). All interviewees gave informed consent. Some signed a virtual form while others gave oral consent during a recorded video call.

## 3.0 Results

### 3.1 Comparison of the economies and health system contexts of Malawi and Zimbabwe

Malawi is a low-income country whose population is largely rural dwelling, with 83% of the country’s 19.4 million people living outside of urban centres. By comparison, Zimbabwe is a lower-middle income country with 68% of the country’s 15.7 million people living in rural areas (Table 1). The gross domestic product (GDP) of these two countries varies greatly, with Malawi being the less well-resourced. Correspondingly the proportion of health expenditure paid ‘out-of-the-pocket’ is higher in Malawi, although it is high in both settings. Notably in both countries, more than half of healthcare expenditure is externally sourced from donors, such that of the two countries, Zimbabwe contributes the smallest proportion of government expenditure towards healthcare (Table 1).

**Table 1:**
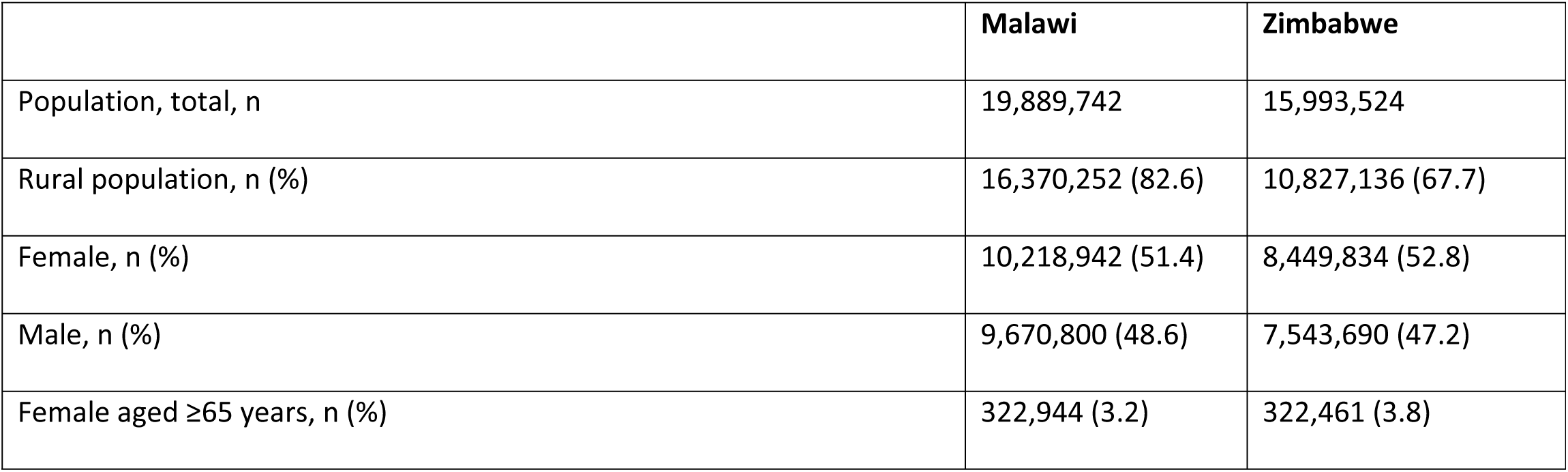
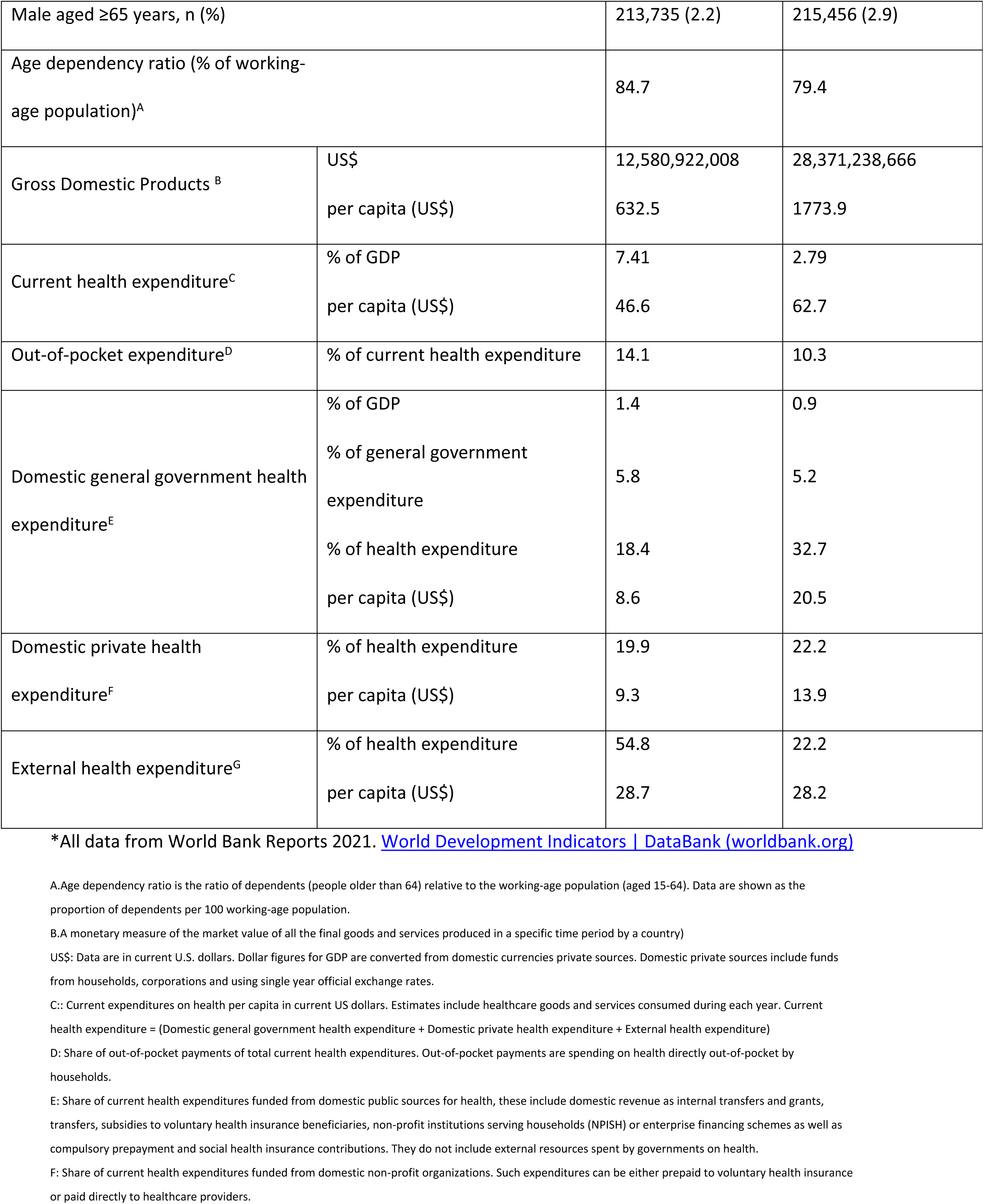
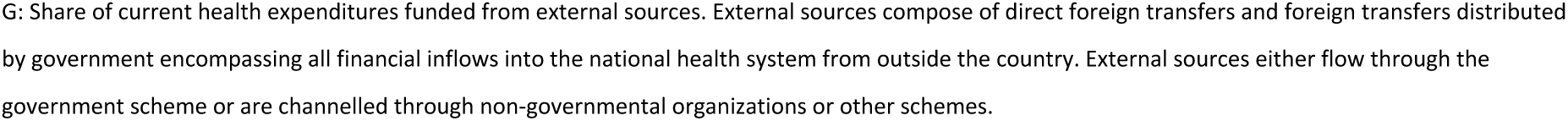
Development Indicators of the healthcare economy in Malawi and Zimbabwe in 2021, according to the World Bank*

The two health systems share characteristics of high public sector dependence, resource scarcity, and verticalization, with some variance in the definitions of the levels of care. In Malawi, in the post-independence era, there was a significant and relatively successful push to establish basic healthcare infrastructure to serve the country’s majority. Most of the healthcare since then has been provided by public funded facilities, which offer free care. Healthcare is also delivered through a network of NGOs, private-for-profit, faith-based-not for profit, informal faith-based healers, traditional healers, who play various roles along the care spectrum. The public facilities are hierarchically ordered and operate on a four-tier service: Community, Primary, Secondary and Tertiary facilities. Chronic disease management (including management of patients with multimorbidity) is provided at all levels of healthcare. At the community level, screening and basic care are provided either in people’s homes, community gatherings, dispensaries or health posts by health surveillance assistants (HSA), community health nurses and community health workers if the resources are available. Healthcare workers in the community refer patients to health centres or community hospitals, which are primary health facilities. Health centres cater to a number of surrounding communities. Health centres refer patients to secondary level district hospitals which are found in 26 of the 28 districts of Malawi. Tertiary hospitals are found regionally and provide most specialised care. Each level of care faces challenges in providing adequate and efficient care.

In Zimbabwe, huge strides were made following independence in 1980 in expanding access to healthcare, including in moving from an urban, curative, and racially-biased health system to one focused on delivering free primary healthcare to underserved communities. Zimbabwe came to boast one of Africa’s strongest health systems, with thriving teaching hospitals and academic sector, excellent laboratory capacity, a well-trained workforce, and a robust primary healthcare infrastructure. The achievements of the 1980s–90s were undone by political instability, structural adjustment (which decreased public spending in favour of privatisation), hyperinflation, the reintroduction of user fees, and the HIV and AIDS epidemic. While Zimbabwe’s system has suffered since the 1990’s, it retains the basic clinic infrastructure and referral system.

Zimbabwe operates on a district health system whose first defined level of care is the primary level. This includes clinics (both public and private), rural hospitals and, in the metropolitan provinces, polyclinics (many of which have satellite units). Community based care, while not a discrete formally defined level of care, is a heterogeneous space that includes community health workers (CHWs) operating out of the primary care facilities, private pharmacies, as well as NGO community support structures for instance the ‘Friendship Bench’ (a community-based peer-provided psychological support model), HIV treatment support services, and grassroots CSOs. The informal sector comprises traditional and faith healers, both politically powerful groups in Zimbabwe with a strong role in healing at the community level but with relatively little formal integration. Zimbabwe’s informal pharmaceutical market, while smaller and more hidden than neighbouring countries (e.g. Malawi), does exist particularly for antibiotics and analgesia and are biased towards the urban centres, with travelling vendors servicing rural areas.

Primary care services in the public sector are nurse-led. However, currently most treatments for NCDs cannot be nurse-initiated, which, given the scarcity of physicians means patients either consulting a private general practitioner/clinic or sourcing a referral to a secondary hospital (defined as Mission, District, Municipal, and most private hospitals), tertiary hospital (only in the rural provinces), or a quaternary (‘central’) hospital (only in metropolitan provinces), depending on what is nearest. In the metropolitan provinces, most often patients are referred to a central hospital for treatment initiation. Stable and uncomplicated patients are then usually referred back to their nearest primary facility for supported self-management (though reviews may still require hospital visits). Unstable and complicated cases are referred to tertiary or quaternary level services where periods of hospitalisation may be required before being referred back to primary facilities for outpatient care. Unlike in Malawi, patients pay a fee for medicines and, in the metropolitan centres, user fees to access health facilities.

Comparison of the care pathways between Malawi and Zimbabwe enabled us to produce a broadly comparable common care pathway encompassing community, primary, secondary, tertiary and quaternary level care (Figure 1). Overall, the kinds of actors comprising these different levels of care - public sector, partner supported, private and informal - are broadly commensurate. The notable difference is that quaternary care only exists in Zimbabwe and, even then, only in the metropolitan provinces, fulfilling essentially the same functions as Malawi’s four tertiary referral hospitals. This common care pathway enables us to discuss core challenges and opportunities in the care of people living with multimorbidity in the following sections.

**Figure 1.**
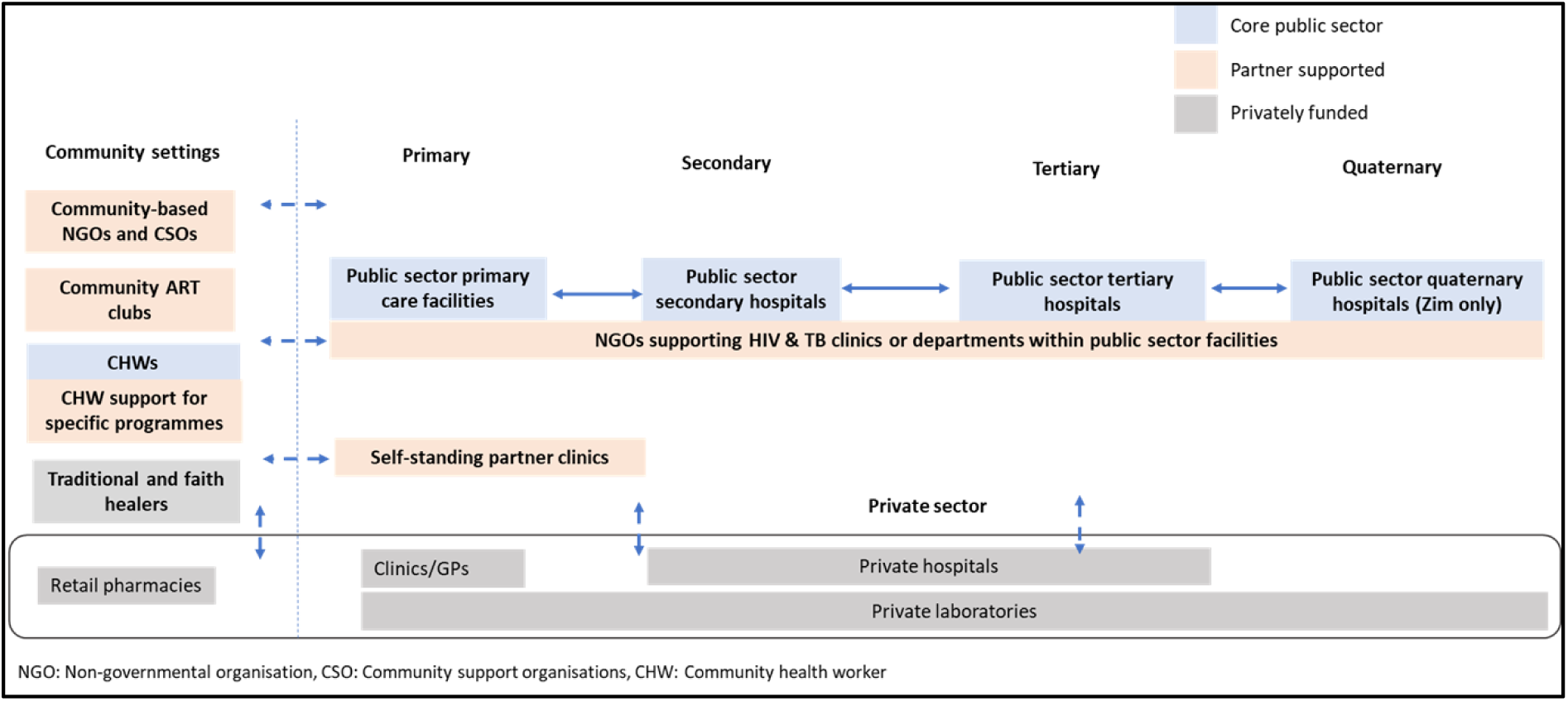
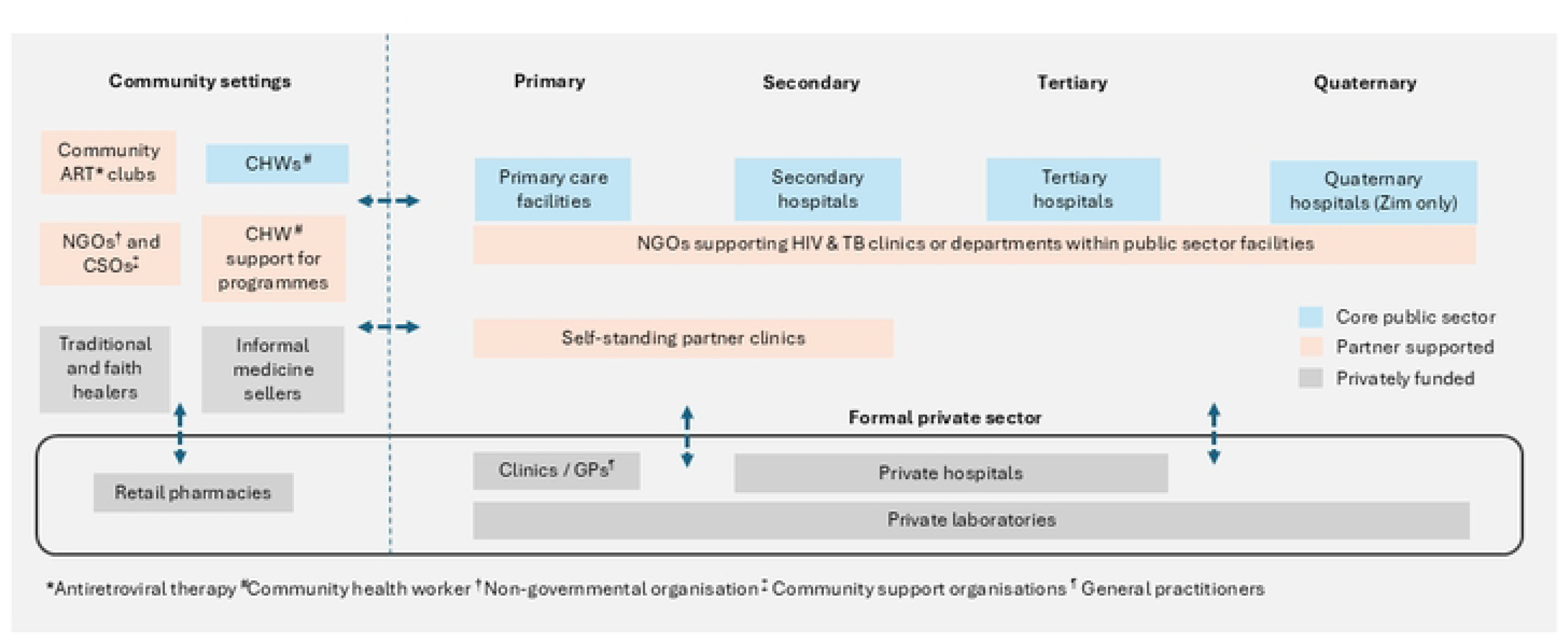
Common care pathway for chronic diseases and multimorbidity in Zimbabwe and Malawi

### 3.2 Challenges and Opportunities in Malawi

A summary of challenges and opportunities for multimorbidity care in Malawi, organised by levels of care, is provided in S3 Table.

#### Community Level

At the community level, challenges within the system complicate multimorbidity management through weak information systems and inadequate human resource, and those present often possess limited skills.

> *“Identification of those diseases is a challenge. If they can at least know that this is hypertension, this is CKD, this could be HIV and then also let them manage them. Not all these conditions could be managed at community level but at least if they can recognise that this is something that needs to be referred to a certain level and also begin the counselling and community sensitization.” (*PM02, Ministry of Local Government)

Policy stakeholders reported that community-based healthcare workers, who often have been trained to prioritise communicable diseases, lacked knowledge of NCDs or multimorbidity. There is also limited supervision by district level providers due to several factors including lack of resources and training. Despite the free public healthcare services, patients often face high out-of-pocket costs to cover transportation to referral centres, which leads to seeking care from traditional healers, who welcome negotiated modes of payment.

> *“The whole reason that it’s traditional for us is because they are available immediately. They are easily accessible within the communities and they have, I should say flexible ways of paying like you can negotiate. When you want to go to the hospital, you need to find transport there. Whereas in the Community, the transport aspect is eliminated, and even if that person be it a, a church person or a traditional healer or a medical man or something, even if they require payment, it can be done in instalments over an extended period of time, which makes sense to the patient”* (PM10, Funder)

Stakeholders reported patients’ lack of knowledge on self-management, dietary restrictions and non-adherence to their medication. They also reported that patients do not understand the chronicity or complexity of their condition, including the need for preventative treatment, and therefore, fail to comply with longer-term prescriptions. Some patients stop taking medication once they are recovered from an acute period of ill health.

> *“Yes and also understanding the disease itself because maybe they will be started on care whilst they are sick, when they get better, they think that’s it. So they don’t understand what it is and the adherence to drugs when they are well is also a challenge.”* (PM02, Ministry of Health)

Support groups coordinated by the resident community health workers (CHWs) seem to facilitate care in terms of ART compliance and raising awareness of NCDs. Most CSOs have focused on HIV/AIDS care and other livelihood programs and not so much on NCD care. For those with NCD support groups, healthcare workers do not always recognize the need for urgent or timely referrals to higher facilities.

#### Primary level

Primary facilities are found both in rural and urban settings and serve as the first entry point for formal health facilities. All primary facilities have at least one trained NCD coordinator (nurse or clinical officer) who oversees NCD related activities within the facility; this helps to prioritise NCD agendas. Although NCD coordinators have helped lobby for resources such as test kits, stakeholders felt that the presence of program-specific coordinators has contributed to siloed care by only focusing on specific diseases.

Primary level care suffers from an inadequate healthcare workforce who receive limited training to adequately care for chronic diseases. Although the referral pathway is clear, there are often delays in patient referral from the primary facilities due to delayed diagnosis and lack of transportation. Diagnostic delays increase the risk of disease severity and complications.

> *“It’s difficult, because those patients usually come when they have a complication. Most of the NCDs are silent, so they don’t know that they are hypertensive, they don’t know that they are living with diabetes. But then let’s say they just come and they are unconscious, or maybe they already have a cerebrovascular accident (stroke).”* (PM02, Ministry of Local Government)

#### Secondary level

Secondary facilities, in the form of district hospitals, are found in almost every district and are the main referral centres for primary facilities. Stakeholders reported inadequate clinical competence among healthcare providers of NCD care compared to HIV services at the secondary level.

> *“I think for me, the biggest challenge is the knowledge gap that is there within health care workers on the conditions. I think they know more on communicable diseases”* (PM09, Ministry of Health)

There is also a lack of guidelines and protocols to inform the management of multimorbidity, and thus judgement is left at the discretion of the clinician, even within the integrated clinics. Patients with multimorbidity require an integrated ‘one-stop-shop’ where all services are provided including pharmacy. In Malawi, integrated NCD clinics through the WHO Package of Essential NCDs (WHO PEN) are common at secondary facilities and have proved useful for people living with multiple NCDs. But HIV services are yet to be integrated in most of the districts (except in Neno where ART and NCD clinics are fully integrated, expanding on the PEN model to include HIV; see Case Study 1). Some secondary care facilities are also piloting an expanded version of the WHO PEN (PEN+). However, comprehensive management is also hindered by lack of physical infrastructure which limits the space available to assess patients in most secondary facilities.

> *“ We have infrastructures that have been there for a long time and have never been maintained and this space is still the old small space.”* (PM01, Ministry of Local Government)

However, other district facilities have been renovated and have adequate space to cater to the number of chronic disease patients that are seen at those facilities (Case Study 1), without patients going back and forth between different departments, or going to facilities on different days to seek care for their individual multimorbidities.

> *“Neno has what we call IC3 integrated chronic care clinic whereby all chronic care conditions (including HIV) are nested under one roof within the same building. You wouldn’t be told to come tomorrow, you are coming today with a condition, you will be supported within the same day and if need be you can be moved within the rooms but without having any hustles or any problems to receive care.”* (PM03, Ministry of Health)

#### Tertiary level

Tertiary facilities (central hospitals) are found regionally and provide specialised care for individual conditions. Inadequate staffing at the tertiary level leads to overwhelmed healthcare workers who have to attend to long queues. Stakeholders felt that this results in poor quality consultations and examinations, where clinical advice was misunderstood, and where only the presenting disease is addressed, and the other comorbidities are ignored; further contributing to diagnostic delays. Patients with multimorbidity visit different departments for different conditions, spending long times at the hospital. Stakeholders reported a lack of guidance and direction to the different specialists the patients are supposed to visit, they are left to explore this on their own.

Stakeholders felt that the upward referral chain is clear but the linkage of patients back to the local referring health facility, so that the patient can continue care, is poor. This leads to patients being tied to the central hospital for ongoing clinical services despite the need to travel long distances and incur associated costs. An incomplete loop (feedback and follow up mechanism) was evident.

> *“In most cases the clinical pathway is not really clear. They do get the BP, the blood pressure checked at the ART clinics, but then now it’s about getting that information to help us also to help them so we might be moving in the right direction, but we need a clear clinical path.”* (PM05, Ministry of Health)

> *“We do actually follow up patients but it’s still posing a challenge because the ART program has very good follow up mechanics to make sure that they are adhering to their medication. But the same people could have diabetes or hypertension in addition to the HIV. So you find that the same kind of system that is there to take care of the ARTs recipients, is not there to cover for these other conditions.”*(PM08, Funder)

### 3.3 Challenges and Opportunities in Zimbabwe

A summary of challenges and opportunities for multimorbidity care in Zimbabwe, organised by levels of care, is provided in S4 Table.

#### Community

Zimbabwe has historically had strong CHW networks from its village health worker programme established in the 1980s. CHWs support primary clinics through health promotion, screening, and follow-up. In theory CHWs are holistically trained to support the management of all conditions managed at primary level; however, funding constraints mean that HIV, TB, malaria, and maternal and child health services occupy most CHWs’ workload. CHWs are overworked and overburdened by the demands of different programmes, and non-partner-supported areas have suffered as a result, particularly NCDs. When asking one district-level stakeholder about NCD outreach, he argued:

> *“Not really, they [NCD outreach activities] are not prioritised that much because at the end of the day it all comes back to funding”* (District Medical Officer, Matabeleland South, Zimbabwe)

This impacts community awareness of NCDs, a lack of equipment with which to screen, lack of ability to follow up patients who have missed visits, and a lack of medicines to provide to patients. Currently much NCD screening in the community is ‘piggybacked’ on donor-funded programmes, and therefore remains patchy.

In terms of multimorbidity, another policymaker spoke of recent guidance to integrate hypertension, diabetes, and mental health into the HIV programme, which includes the regular screening (including by CHWs) of these conditions among people living with HIV. This could present an opportunity to expand NCD community outreach/ follow-up; however, this initiative remains in its early-stages, with shortages of equipment and medicines and, those that do exist being restricted to people with HIV. For the majority, community-based care for NCDs and multimorbidity (if conditions are even known to clients/facilities) comprises buying medicines over the counter, consulting traditional and faith healers, and using home remedies. This isolation of NCD treatment seeking is compounded by a lack of peer support groups, which have enjoyed varying degrees of success within the HIV programme. There is ongoing research on peer support groups for NCD care including self-monitoring and bulk purchasing of medicines, but currently such activities remain contained within small-scale research settings.

#### Primary level

Primary care has historically been a strength in Zimbabwe. While this backbone remains, the superimposition of vertical programmes for HIV, TB, and malaria has resulted in a stark disparity in care for these conditions relative to NCDs. Within public sector primary care facilities, services for HIV and TB are free, decentralised and often delivered through dedicated HIV clinics within primary healthcare. Certain independent partner-funded clinics are even able to extend this free package to include ‘one-stop-shop’ care for NCDs tailored to the life course (see Case Study 2). While there is the momentum to integrate NCDs into HIV care in the mainstream public sector, currently the management of clients with HIV and NCDs will generally be split between HIV clinics and general outpatient departments (OPDs), which means multiple queues, user and medicines fees, recurrent referrals to pharmacies for medicines given the persistent stockouts, and referrals to hospitals for treatment initiation and reviews. Weak information systems make it extremely challenging to follow-up NCDs and to understand population-level health statistics.

In parallel to NCD integration within the HIV programme, the WHO PEN and PEN+ programme is currently being piloted in several districts to enable decentralisation and integrated management of key NCDs at primary level. Much hope has been pinned to the PEN + programme among policymakers with a focus on NCDs:

> *“In PEN+ doctors are enrolled to do visits per month to primary healthcare so as to cut down costs for patients. Medication that would have been prescribed by the specialist is then made available at the local clinic so that collection is done at clinic level rather than the patient having to travel to get medication. The monitoring is also intensified via community linkages through community health workers and the family involved to make sure the patient adheres to the medication”*. (Policymaker, MoHCC, Harare)

While this remains a key programme for expanding care for NCDs beyond HIV, PEN faces continuing resource challenges, and many costs, including user fees and medicines fees, are still shouldered by patients. The other possible recourse for NCD and multimorbidity care is found in the private sector, where family physicians with strong generalist skills can be accessed as well as specialists for different NCDs (some requiring travel to South Africa). However, for the majority private care was unaffordable. Even those who may initially afford it would often have their resources so depleted that they must switch to access the public sector and its challenges.

#### Secondary and tertiary level

District and provincial hospitals, only located within the rural provinces, generally have physicians available, who can make routine diagnoses, and initiate treatments of most common NCDs, with better linkage to primary care services, compared to Malawi. Inpatient and certain specialist services were also available particularly at the tertiary level, though this was highly variable with many posts vacant (e.g. one provincial hospital had only one general surgeon, one orthopaedic surgeon, and one obstetrician and gynaecologist). The same resource challenges and disparities in NCD care relative to HIV were equally experienced by the hospitals. Challenges commonly cited included staff shortages and attrition – clinicians often leaving as soon as the opportunity arose – medicine stockouts, and shortages of beds, equipment, commodities, laboratory infrastructure and reagents. Additionally, beyond the few hospitals currently implementing PEN or PEN+, patients with NCDs had to travel significant distances to see the doctor:

> *“We need to schedule a day for the doctor to come and see patients alongside a pharmacist. Patients are struggling to go to [District Hospital] and so they end up not going”* (Primary care nurse, Matabeleland South)

As a result of these challenges, often by the time a patient reaches a district or provincial hospital they are extremely sick with complications, requiring inpatient care. Such inpatient stays can be extremely expensive for patients and families because of the higher user fees relative to primary care and likely, in any case, require onward referral given the relatively few speciality services available, not to mention the paucity of even basic resources at the hospital. Transporting very sick, often critical patients to higher levels of care in the metropolitan centres of Harare and Bulawayo was reported as a severe drain on hospital resources, and, ironically, one of the reasons why any spare funds went on transport rather than purchasing out-of-stock NCD medicines.

#### Quaternary level

The central hospitals, located in the urban metropoles, provide the highest level of care, with most specialised services being broadly comparable to Malawi’s tertiary referral hospitals. One of the main challenges experienced in both Harare and Bulawayo was that patients were choosing to ‘bypass’ local clinics due to the unavailability of resource at the primary level, and instead present directly to hospitals, which disrupted the referral pyramid and overwhelmed the central hospital casualty and outpatient departments:

> *“We have historically had strong primary healthcare, but we’re riding on what used to be. ‘Level 1’ doesn’t exist as much as it used to, as we have people coming to UBH first point of care, so the levels of care aren’t as defined as previously. They’re going straight to the specialist especially for NCDs, instead what the doctor and nurse used to be able to do community based”* (Policymaker, Workshop proceedings, Harare)

Aside from donor-supported HIV centres of excellence located at most central hospitals, the hospitals were also seriously under-resourced (and even the Centres of Excellence faced resource challenges for patients’ NCD comorbidities). Challenges included staff shortages and attrition; a lack of in-country sub-specialist training (students would have to train overseas, self-funded); lack of equipment and infrastructure (e.g., running water, reliable electricity supply) and specialist treatments to actually practice specialty medicine (e.g., limited dialysis capacity, cancer treatment, not all surgeries that need to be performed can be). The unavailability of laboratory investigations meant inpatients and outpatients were often referred to off-site private laboratories, and these tests could cost many hundreds of dollars.

Patients with NCDs and multimorbidity experienced particular challenges accessing quaternary care. Despite growing emphasis on multidisciplinary and improved communication between different specialists, in reality patients with multimorbidity would have to attend appointments for different conditions on different days, which was a considerable burden on time and resources. Because of the challenges of access and delays seeking care, many patients were critically ill by the time they arrived (often having been transported from secondary and tertiary hospitals) and would require prolonged inpatient stays that were paid out of pocket and extremely expensive, proving catastrophic for most. Respondents reported increasing emphasis at Ministry level on strengthening specialist capacity at the central hospitals to attend to the current gaps and challenges in services. However, others noted that this emphasis was at the expense of strengthening generalist capacity, preventative medicine and public health community based which would enable many of the conditions currently being seen at central level being prevented or managed in communities to unburden the central Hospitals.

##### Box 1: Integrated Chronic Care Clinic, Malawi

**Case Study 1: Integrated Chronic Care Clinic, Malawi**

The integrated chronic care clinic (IC3) model was established in 2015 by introducing the WHO Package of Essential Non-Communicable Diseases (PEN) to an existing HIV programme and is implemented by Abwenzi Pa Za Umoyo/Partners In Health (APZU/PIH) through and with the Ministry of Health in Neno. The IC3 takes care of patients with chronic illnesses enrolled in longitudinal care.

The IC3 model begins with identifying and following up individuals at facility-level and follows them up at household-level using a team of community volunteers, community health workers (CHWs). CHWs complete monthly household visits aimed at screening (blood pressure, random blood glucose), linkage to care and “shoulder-to-shoulder” patient accompaniment to facilities. At community level, active case finding initiatives are routinely completed through what is termed integrated Screening for Health and Referral at the Community (SHARC) activities. Similar to SHARC, all facilities have Screening for Health and Referral at the Facility (SHARF) initiatives within the IC3 programming. All patients that screen positive for a condition are then linked to care through the CHWs and digitally documented through CommCare digital app for follow up at the nearest facilities.

As of June 2024, 12% of patients with NCD were also on HIV treatment. One-year retention for NCDs was 84% and for HIV was 95%, with mortality rates of less than 1 percent for both NCD and HIV cohorts. Clinical outcomes for HIV-NCD comorbid patients showed good virological control ranging from 85 to 95%. Clinical outcomes and retention in care are favourable, suggesting that integration of chronic disease care at the primary care level is a feasible way forward for settings with a dual burden of HIV and chronic NCDs.

##### Box 2: Newlands Clinic, Zimbabwe

**Case Study 2: Newlands Clinic, Zimbabwe**

Newlands Clinic, in Harare, Zimbabwe, established in 2004, is a non-governmental organisation operated by the Ruedi Luethy Foundation. In partnership with the Zimbabwean Ministry of Health and Child Care, the clinic provides comprehensive, nurse-led, outpatient treatment and care for 7,800 people living with HIV. Now, 35% patients are 50 years or older, growing from 15% in 2013. To provide holistic care to the ≥65-year-old population a novel integrated HIV-geriatric clinic was set-up. Older people are seen with their caregiver, and a structured nurse-led comprehensive geriatric assessment is performed. This clinic aims to provide annual assessments and management of multimorbidity, developing personalized care plans to optimize disease management, and functional ability.

Among the first 100 patients assessed (2022–23), whilst 94% had undetectable viral loads (<50 copies/ml), multimorbidity was common with a median number of morbidities per person of 3 (IQR 2-4), most were previously undiagnosed; hypertension, diabetes, chronic kidney disease, visual and hearing impairments, and depression/anxiety were the most common. Cardiovascular risk was high in 43%, and statins were initiated. Overall, 86% received a new clinical intervention and 76% received at least one new diagnosis. Feedback from patients was very positive. This case study shows, albeit with appropriate financial support, it is feasible to set-up and run an integrated HIV-geriatric clinic in Southern Africa, and when looking beyond HIV management, multimorbidity is common, addressable and the management is valued.

## 4.0 Discussion

Multimorbidity is a growing public health concern in Malawi and Zimbabwe, as in many sub-Saharan African countries. In this study, we mapped patient journeys in care seeking for multimorbidity in Malawi and Zimbabwe to identify key stages and steps that determine care access along the care pathway, as well as challenges and facilitators in accessing care. In both countries, services are provided at a variety of publicly funded, faith-based, not-for-profit and for-profit organisations; services are fragmented and there are highly pluralistic health systems. Our study highlights country-specific contextual, logistical and human resource barriers that affect care delivery, including challenges to the integration of HIV and NCDs, as well as opportunities for targeted interventions.

Key commonalities across these neighbouring countries can form the basis of further cross-country research and health systems intervention development.

Our results paint a very different picture from chronic care journeys in higher-income settings, for instance Maas et al., showed that in the Netherlands patients living with chronic conditions, whether one or multiple, seek care in a “logical” loop between self-initiated management at home and seeking facility-based care.(22) In the majority of lower-resource settings, such as ours, theoretically there may be a loop, but this is broken. As a consequence, individuals are often forced to “skip” referral points to access higher levels directly, whilst other patients are referred back and forth between their nearest facility and a higher referral centre. The reasons for this heterogeneous care seeking are multiple such as the unpredictability and progression of chronic conditions and the lack of preparedness of the health system at all levels to manage such complexities. Indeed inadequate staff trained available to manage NCDs, especially at primary facilities, has been reported in other sub-Saharan African countries, and contributes to the “system bypass”, and thus can lead to the back and forth between levels of care.(23,26) Our findings are consistent with other studies that have highlighted non-linearity and turbulence of care pathways, arising due to numerous, interlinked challenges at each level of care, such as lack of human resources and skills, lack of standardized guidelines to inform care, and patient information resources that might otherwise promote self management.(5, 24, 25, 27)

With these care patterns and health-seeking behaviours, multimorbid patients incur high out of pocket expenditures due to visiting multiple health facilities and departments. And as a result of limited medical resources, which are mostly observed in public facilities, they are forced to buy medications and seek treatment in private facilities.(28) In most of LMICs, a large proportion of people are uninsured, meaning individuals incur catastrophic costs, pushing people into poverty.(29) Managing these patients increases cost due to prolonged and multiple consultations and multiple prescriptions and polypharmacy. This highlights the need for comprehensive and cross-department integrated care models. (30)

Our study captured several initiatives to integrate chronic care services in Malawi and Zimbabwe, as is increasingly being explored in several countries across sub-Saharan Africa.(14) As has been the experience in other countries, hypertension and diabetes screening have been integrated in many public sector HIV clinics, operating as an expansion of the vertical programme for those living with HIV.(31, 32) In some self-standing partner-funded contexts, for instance Newlands Clinic in Zimbabwe, this has resulted in extremely high-quality integrated care, inclusive of free medicines for NCDs and may, indeed, constitute among the most person-centred approaches available on the continent. However, outside these well-funded contexts, often screening is not matched with treatment. In both Malawi and Zimbabwe, medication prescription for these conditions is largely not done within these integrated HIV clinics. This means that despite integration, patients visit multiple clinics to seek care for all their ailments. Even where outpatient clinics have been integrated, lack of diagnostic equipment, poorly trained healthcare workers and frequent drug stockout undermine clinical management. Delays in diagnosis due to equipment stockouts have been reported in other sub-Saharan African countries.(33) Delayed diagnosis coupled with lack of NCD medication can threaten the management of multimorbidity and further complicate disease burden, which overall effects integration.

Outside of the HIV platform, stakeholders in both countries reported moving towards PEN and PEN+ models. Unlike in Malawi where PEN + had been fully implemented at two secondary facilities by 2020, WHO PEN+ models are being piloted at primary facilities in Zimbabwe.(34) Already, trainers from Malawi have been part of the team training the new PEN+ practitioners in Zimbabwe.(38) The IC3 model applied in Neno district could form a common frame of reference for optimising cross-country learning and collaboration to support integrated chronic care in both Zimbabwe and Malawi. The integration of chronic diseases management and implementation through an expanded PEN+ model, integrated with HIV, could lead to leveraging historically earmarked HIV funding while cost saving by consolidating services and reducing the need for patients to attend multiple facilities while also accessing existing health resources and infrastructures. Concurring with other studies, integration of care may help reduce health care access disparities especially in primary health care facilities, cost and improve outcomes.(39) This is important given that HIV funding is going to continue dropping in sub-Saharan Africa in the build-up to 2030 and the shift towards ‘maintenance’ and following unprecedented funding cuts from the United States Government.

Our study joins a chorus of critical global health scholarship that highlights that external funders skew the health agenda, not just in terms of funding but also in terms of their priorities, such that focus on TB, HIV, malaria has distracted from NCDs for years.(40, 41) Funders involvement in driving the health agenda is a well reported phenomena.(42) Shifting to a chronic disease focus (outside of HIV) will require multi-sectoral involvement and funders may need to rescind some power enable integration of NCD management.(43) It may also require governments to take ownership and leverage HIV program successes, especially in light of major funding withdrawal within the HIV sector. Our stakeholders in both Malawi and Zimbabwe reported that access to antiretroviral therapies (ARTs) is good, which means that a functional supply chain for medicines distribution is possible and effective, yet access to NCD medicines is very poor. Inequalities in the medicine supply chain between HIV and NCDs has been attributed to vertical funding.(44) Even within integrated HIV and NCD clinics, HIV is still seen as the core condition onto which NCDs are piggybacked.

Multimorbidity affects multiple health domains and should force health systems to consider the whole person in their social context, which is where the transformative potential has been argued to lie.(45) Many reviews of integrated services have been too restrictive in their consideration of the scope of multimorbidity and have tended to reproduce, rather than forge a pathway beyond, a disease-centred perspective. Often, epidemiological studies and health system interventions focus on just a couple of different conditions, frequently excluding conditions and associated needs that are not only common, for instance, mental health conditions, but are also highly important to patients.(46, 47) Wong et al, highlighted that communicable diseases such as tuberculosis can also chronically affect individuals and should be part of the multimorbidity discussion.(48) An integrated multimorbidity approach may be a tool to deliver people-centred care beyond the lens of their disease index.(49)

This paper is founded on north-south and south-south partnerships that form part of the African multimorbidity alliance. The Academy of Medical Science has highlighted the importance of such partnerships in the race to curb the impact of multimorbidity(6, 50). Triangulation of our data from various sources is a strength, providing insights from a wide group of policymakers, enabling data corroboration. We, as authors, are from multiple countries which helped our research teams (and organisations) share knowledge and learn from each other’s strengths and weaknesses. Such collaborations can foster conversations around systems thinking across the countries so as to not “reinvent the wheel”.

However, our study has limitations. Despite extracting data from various sources, there may be variations in the patient journey that are unique to either a specific group of patients, or certain chronic diseases which we did not capture. Our study also did not include patient perspectives as users of the system, nor did we seek perspectives from healthcare workers who manage the patients at the different levels of care. We recommend that future research should gain perspectives of people living with multimorbidity and those who provide clinical care. We have made further recommendations in box 3.

## 5.0 Conclusion

Our multi-country study shows similar patterns in how patients with multimorbidity flow through health services. In both settings, patients often seek care in a pluralistic manner cutting across various service providers. Integrated multimorbidity care has been tacitly framed as HIV+ NCDs (in some parts of Malawi) or only selected NCDs in both countries, systems driven by donor-funded vertical programs have pushed HIV services to be exceptional, leaving NCD services “orphaned”, and some NCDs such as mental health neglected. Multimorbidity care in both settings is often hindered by inadequate and poorly skilled human resource, frequent drug stockouts and lack of diagnostic capacity. Expanding the scope of multimorbidity beyond HIV and selected NCDs offers a much-needed people-centred approach, which multiple stakeholders agree is the best way to care for people living with multimorbidity.

## Data Availability

All data generated or analysed during this study are included in this published article [and its supplementary information files]. Anonymised copies of original transcripts can be shared upon reasonable request.

## Acknowledgements

We would like to acknowledge Dr Edna Bosire, Dr Myrna Van Pinxteren and the members of the Africa Multimorbidity Alliance for their support during the conceptualisation and the writing stage. We acknowledge the efforts and support of the grant holders within the Multilink and KnowM consortiums.

## Authors contribution

All authors conceptualised the study. GTB and JD collected and analysed the interviews. CLG, BM and BDK collated the case studies. FL and JD collated the figures. GTB and MJG drafted the first manuscript. All authors have read, critically reviewed and approved the final manuscript

### Box 3: Recommendations for strengthening pathways of health care for people living with multimorbidity in low-income settings.

**Community healthcare level**

1. Community level information systems for early NCD screening, diagnosis and treatment, and NCD awareness including development of community health prevention services
2. Community health workers have adequate information and competencies on multimorbidity management through trainings and supervision
3. Patient self-management education after health facility care to enhance lifestyle modification and adherence to medication.
4. Traditional healers are incorporated as key stakeholders along the care pathway in management of NCDs for screening and prompt referral to primary health care.

**Primary healthcare level**

1. Healthcare workers are competent and equipped to provide integrated disease screening, diagnosis and ongoing management of patients with multimorbidity
2. Routine supportive supervision visits from senior clinicians in district hospital would enhance competence and confidence among health workers in primary healthcare facilities
3. Basic but essential medicine and diagnostic equipment are available, functional and adequate for screening, diagnosing and treating patients without overburdening patients with costs
4. Referral systems are developed, strengthened and are responsive to avoid diagnostic and care delays, preventing secondary healthcare being overwhelmed by disease complications e.g. stroke

**Secondary healthcare level**

1. Healthcare workers are competent and equipped to provide integrated disease screening and ongoing management of patients with multimorbidity
2. Supportive and adequate infrastructure provides enough space for confidential patient consultations and well-equipped laboratories for efficient sample processing.
3. Learn and leverage existing programmes to inform expansion to other parts of the country
4. Essential medicine and diagnostic equipment are available, functional and adequate for screening, diagnosing and treating patients without overburdening patients with costs.

**Tertiary and Quaternary healthcare level**

1. An adequate number of specialists to provide care as well as undertake district visits, with clinical competence in providing NCD services
2. Closing the loop of the referral system would ensure that patients are referred back, with clear communication, to primary care for ongoing community-based care and support.
3. Integrated management of multimorbidity that prevents patients from having multiple visits to the hospital (high cost) with the complexity of navigating multiple departments
4. By strengthening primary and secondary level care, patient loads will be lessened in tertiary hospitals, thereby promoting a more sustainable pathway of care.

## Supplementary materials

### S1 Figure

S1 Table:Table summarising the included policy documents

S2 Table: Table of healthcare providers

S3 Table: Table showing the challenges and opportunities in Malawi

S4 Table: Table showing the challenges and opportunities in Zimbabwe

## References

1. Pearson-Stuttard J, Ezzati M, Gregg EW. Multimorbidity—a defining challenge for health systems. The Lancet Public Health: Elsevier Ltd; 2019. p. e599–e600.

2. Souza DLB, Oliveras-Fabregas A, Minobes-Molina E, de Camargo Cancela M, Galbany-Estragués P, Jerez-Roig J. Trends of multimorbidity in 15 European countries: a population-based study in community-dwelling adults aged 50 and over. BMC Public Health. 2021;21(1).

3. Guthrie B, Barnett K, Mercer SW, Norbury M, Watt G, Wyke S. Epidemiology of multimorbidity and implications for health care, research, and medical education: a cross-sectional study. Lancet. 2012;380:37–43.

4. Spencer SA, Yongolo NM, Simiyu IG, Sawe HR, Dark P, Dula D, et al. The Burden of Multimorbidity-Associated Acute Care Hospital Admissions in Malawi and Tanzania: A Prospective Multicentre Cohort Study. 2024.

5. Chikumbu EF, Bunn C, Kasenda S, Dube A, Phiri-Makwakwa E, Jani BD, et al. Experiences of multimorbidity in urban and rural Malawi: An interview study of burdens of treatment and lack of treatment. PLOS Global Public Health. 2022;2(3 March):e0000139–e.

6. The Academy of Medical S. Improving the prevention and management of multimorbidity in sub-Saharan Africa. Johannesburg; 2019 2019/9//.

7. Spencer S, Rylance J, Quint J, Gordon S, Dark P, Morton B. Use of hospital services by patients with chronic conditions in sub-Saharan Africa: a systematic review and meta-analysis. Bulletin of the World Health Organization. 2023;101(09):558–70G.

8. Basto-Abreu A, Barrientos-Gutierrez T, Wade AN, Oliveira de Melo D, Semeão de Souza AS, Nunes BP, et al. Multimorbidity matters in low and middle-income countries. Journal of Multimorbidity and Comorbidity. 2022;12:263355652211060-.

9. Glynn LG, Valderas JM, Healy P, Burke E, Newell J, Gillespie P, et al. The prevalence of multimorbidity in primary care and its effect on health care utilization and cost. Family Practice. 2011;28(5):516–23.

10. Kivuyo S, Birungi J, Okebe J, Wang D, Ramaiya K, Ainan S, et al. Integrated management of HIV, diabetes, and hypertension in sub-Saharan Africa (INTE-AFRICA): a pragmatic cluster-randomised, controlled trial. The Lancet. 2023;402(10409):1241–50.

11. Barnett K, Mercer SW, Norbury M, Watt G, Wyke S, Guthrie B. Epidemiology of multimorbidity and implications for health care, research, and medical education: A cross-sectional study. The Lancet. 2012;380(9836):37–43.

12. Salisbury C. Multimorbidity: redesigning health care for people who use it. The Lancet. 2012;380(9836):7–9.

13. Collini P, Mawson RL. A new era of HIV care for age-associated multimorbidity. Current Opinion in Infectious Diseases. 2023;36(1):9–14.

14. McCombe G, Lim J, Hout MCV, Lazarus JV, Bachmann M, Jaffar S, et al. Integrating Care for Diabetes and Hypertension with HIV Care in Sub-Saharan Africa: A Scoping Review. International Journal of Integrated Care. 2022;22(1).

15. Matanje Mwagomba BL, Ameh S, Bongomin P, Juma PA, MacKenzie RKKC, Lukhele N, et al. Opportunities and challenges for evidence-informed HIV-noncommunicable disease integrated care policies and programs lessons from Malawi, South Africa, Swaziland and Kenya. AIDS. 2018;32:S21–S32.

16. Kasomekela N. Malawi PEN-Plus Operational Plan 2021. 2021.

17. Banda GT, Bosire E, Bunn C, Chandler CIR, Chikumbu E, Chiwanda J, et al. Multimorbidity research in Sub-Saharan Africa: Proceedings of an interdisciplinary workshop. Wellcome Open Research. 2023;8:110-.

18. Jones J, Hunter D. Qualitative Research: Consensus methods for medical and health services research. BMJ. 1995;311(7001):376–80.

19. Cathy W. Multilink Consortium.

20. Zim T. KnowM. Thru2021.

21. Dixon J, Dhodho E, Mundoga F, Webb K, Chimberengwa P, Mhlanga T, et al. Assembling the Challenge of Multimorbidity in Zimbabwe: A Participatory Ethnographic Study. 2024.

22. Maas VK, Dibbets FH, Peters VJT, Meijboom BR, van Bijnen D. The never-ending patient journey of chronically ill patients: A qualitative case study on touchpoints in relation to patient-centered care. PLoS ONE. 2023;18(5 May).

23. Morgan SA, Eyles C, Roderick PJ, Adongo PB, Hill AG. Women living with multi-morbidity in the Greater Accra Region of Ghana: a qualitative study guided by the Cumulative Complexity Model. Journal of Biosocial Science. 2019;51(4):562–77.

24. Eyowas FA, Schneider M, Alemu S, Getahun FA. Experience of living with multimorbidity and health workers perspectives on the organization of health services for people living with multiple chronic conditions in Bahir Dar, northwest Ethiopia: a qualitative study. BMC Health Services Research. 2023;23(1):232-.

25. Matima R, Murphy K, Levitt NS, BeLue R, Oni T. A qualitative study on the experiences and perspectives of public sector patients in Cape Town in managing the workload of demands of HIV and type 2 diabetes multimorbidity. PLoS ONE. 2018;13(3).

26. Badacho AS, Mahomed OH. Facilitators and barriers to integration of noncommunicable diseases with HIV care at primary health care in Ethiopia: a qualitative analysis using CFIR. Frontiers in Public Health. 2023;11:1247121-.

27. Mitambo C, Khan S, Matanje-Mwagomba BL, Kachimanga C, Wroe E, Segula D, et al. Improving the screening and treatment of hypertension in people living with HIV: An evidence-based policy brief by Malawi’s knowledge translation platform. Malawi Medical Journal. 2017;29(2):224–8.

28. Bähler C, Huber CA, Brüngger B, Reich O. Multimorbidity, health care utilization and costs in an elderly community-dwelling population: a claims data based observational study. BMC Health Services Research. 2015;15(1):23-.

29. Wagstaff A, van Doorslaer E. Catastrophe and impoverishment in paying for health care: With applications to Vietnam 1993-1998. Health Economics. 2003;12(11):921–33.

30. Wang L, Si L, Cocker F, Palmer AJ, Sanderson K. A Systematic Review of Cost-of-Illness Studies of Multimorbidity. Applied Health Economics and Health Policy. 2018;16(1):15–29.

31. Shayo EH, Kivuyo S, Seeley J, Bukenya D, Karoli P, Mfinanga SG, et al. The acceptability of integrated healthcare services for HIV and non-communicable diseases: experiences from patients and healthcare workers in Tanzania. BMC Health Services Research. 2022;22(1):655-.

32. Duffy M, Ojikutu B, Andrian S, Sohng E, Minior T, Hirschhorn LR. Non-communicable diseases and HIV care and treatment: models of integrated service delivery. Tropical Medicine and International Health: Blackwell Publishing Ltd; 2017. p. 926–37.

33. Mulupi S, Ayakaka I, Tolhurst R, Kozak N, Shayo EH, Abdalla E, et al. What are the barriers to the diagnosis and management of chronic respiratory disease in sub-Saharan Africa? A qualitative study with healthcare workers, national and regional policy stakeholders in five countries. BMJ Open. 2022;12(7):e052105–e.

34. Kasambara A, Kamndaya MS, Masangwi SJ, Mulaga A. Non-communicable diseases and HIV/AIDS burden by socio-demographic characteristics in Malawi. Journal of Global Health Reports. 2023;7:e2023080–e.

35. Wroe EB, Kalanga N, Dunbar EL, Nazimera L, Price NF, Shah A, et al. Expanding access to non-communicable disease care in rural Malawi: outcomes from a retrospective cohort in an integrated NCD–HIV model. BMJ Open. 2020;10(10):e036836–e.

36. Wroe EB, Kalanga N, Mailosi B, Mwalwanda S, Kachimanga C, Nyangulu K, et al. Leveraging HIV platforms to work toward comprehensive primary care in rural Malawi: the Integrated Chronic Care Clinic. Healthcare. 2015;3(4):270–6.

37. Partners In Health.

38. Network NP. PEN-Plus Providers in Zimbabwe Receive Heart Failure and Echocardiography Training. 2023.

39. Rocks S, Berntson D, Gil-Salmerón A, Kadu M, Ehrenberg N, Stein V, et al. Cost and effects of integrated care: a systematic literature review and meta-analysis. European Journal of Health Economics. 2020;21(8):1211–21.

40. Khan MS, Meghani A, Liverani M, Roychowdhury I, Parkhurst J. How do external donors influence national health policy processes? Experiences of domestic policy actors in Cambodia and Pakistan. Health Policy and Planning. 2018;33(2):215–23.

41. Gautier L, Ridde V. Health financing policies in Sub-Saharan Africa: government ownership or donors’ influence? A scoping review of policymaking processes. Global Health Research and Policy. 2017;2(1):23-.

42. Mwisongo A, Nabyonga-Orem J, Yao T, Dovlo D. The role of power in health policy dialogues: lessons from African countries. BMC Health Services Research. 2016;16(S4):213-.

43. Garrib A, Birungi J, Lesikari S, Namakoola I, Njim T, Cuevas L, et al. Integrated care for human immunodeficiency virus, diabetes and hypertension in Africa. Transactions of the Royal Society of Tropical Medicine and Hygiene: Oxford University Press; 2019. p. 808–11.

44. Mussa AH, Pfeiffer J, Gloyd SS, Sherr K. Vertical funding, non-governmental organizations, and health system strengthening: Perspectives of public sector health workers in Mozambique. Human Resources for Health. 2013;11(1):11-.

45. Dixon J, Morton B, Nkhata MJ, Silman A, Simiyu IG, Spencer SA, et al. Interdisciplinary perspectives on multimorbidity in Africa: Developing an expanded conceptual model. PLOS Global Public Health. 2024;4(7):e0003434-e.

46. Price AJ, Jobe M, Sekitoleko I, Crampin AC, Prentice AM, Seeley J, et al. Epidemiology of multimorbidity in low-income countries of sub-Saharan Africa: Findings from four population cohorts. PLOS Global Public Health. 2023;3(12):e0002677-e.

47. Price AJ, Crampin AC, Amberbir A, Kayuni-Chihana N, Musicha C, Tafatatha T, et al. Prevalence of obesity, hypertension, and diabetes, and cascade of care in sub-Saharan Africa: a cross-sectional, population-based study in rural and urban Malawi. The Lancet Diabetes and Endocrinology. 2018;6(3):208–22.

48. Wong EB, Olivier S, Gunda R, Koole O, Surujdeen A, Gareta D, et al. Convergence of infectious and non-communicable disease epidemics in rural South Africa: a cross-sectional, population-based multimorbidity study. The Lancet Global Health. 2021;9(7):e967–e76.

49. Secretariat. Framework on integrated, people-centred health services Report by the Secretariat. 2016 2016/4//.

50. The Academy of Medical S. Multimorbidity: a priority for global health research. London; 2018.

